# Association between a computerized, Self-administered cognitive assessment and phosphorylated Tau in Alzheimer’s Disease

**DOI:** 10.1101/2024.11.23.24317846

**Authors:** Mina Aghaei, Zahra Vahabi, Mohammad Hadi Modarres, Fatemeh Ebrahiminia, Seyed-Mahdi Khaligh-Razavi

## Abstract

**Introduction:** Alzheimer’s Disease (AD) is the leading cause of dementia, accounting for 80% of cases globally. With an aging population, AD poses a growing concern due to its increasing mortality rate and associated healthcare costs. Early diagnosis or prediction of AD is crucial for managing and potentially slowing its progression, particularly with the advent of disease-modifying therapies.

**Methods:** This study involved a total of 60 participants: 15 with mild AD, 23 with mild cognitive impairment (MCI), and 22 healthy controls, comprising both males and females aged 50–85 years. Data collection included blood samples, the Montreal Cognitive Assessment (MoCA), and the Integrated Cognitive Assessment (ICA). Serum phosphorylated tau181 (p-tau181) levels were measured using enzyme-linked immunosorbent assay (ELISA) kits. Linear regression models were applied to predict serum biomarker levels based on the ICA index, demographic data, and APOE ε4 status.

**Aims and Objectives:** The primary aim was to examine the association between ICA scores and blood-based p-tau181 levels in individual with MCI, mild AD, and elderly healthy controls. Additionally, the study compared the correlation of ICA and MoCA with APOE ε4 status.

**Findings:** The study found that the ICA can significantly differentiate between diagnostic groups and that elevated serum p-tau181 levels are associated with cognitive decline, as measured by the ICA. Additionally, a significant correlation was observed between APOE ε4 status and cognitive decline, but not between APOE ε4 status and serum p-tau181 levels. Using a model that incorporates the ICA, demographic data, and APOE ε4 status, we were able to modestly predict serum p-tau181 levels, indicating the potential of combining cognitive assessment with biological markers for AD prediction.

**Discussion:** The findings suggest that ICA is a valuable tool in detecting cognitive decline associated with AD and correlates with increased p-tau181 levels. The significant correlation between APOE ε4 status and cognitive decline highlights its potential role in risk assessment, independent of p-tau181 levels. The study supports the use of a computerized, self-administered cognitive assessment in conjunction with blood-based biomarkers for early AD detection. Limitations include the small sample size and cross-sectional design, which restricts causal inference and longitudinal analysis. Future research should focus on larger, longitudinal studies to further validate these associations and their implications for early diagnosis and monitoring of AD progression.

## Introduction

Alzheimer’s Disease (AD) is the most common cause of dementia, accounting for 80 percent of cases worldwide. With the aging population, it has become a significant concern with increasing mortality rate and total cost associated with the disease; however, early diagnosis or prediction of the disease can aid in managing and preventing its progression, especially with the recent development and FDA approvals of disease-modifying therapies (1,2).

Cognitive impairment is a key characteristic of dementia. As a result, various cognitive assessments have been developed to screen for cognitive impairment, differentiate its causes, evaluate the severity of the condition, and monitor disease progression. The Integrated Cognitive Assessment (ICA) is an AI-powered digital biomarker designed to evaluate cognition. It focuses on cognitive domains and brain regions typically impacted in the early stages of cognitive disorders like dementia, ideally detecting issues before memory symptoms appear (3).

Previous studies have shown a significant association between ICA index and the thickness of the cortex in key brain areas affected by tau-pathology, including Para-hippocamp in dementia (4). Additionally, ICA index could better differentiate healthy controls from MCI or mild-AD compared to the hippocampal volume, suggesting that poor performance in ICA precedes the impairment of hippocampal volume (5).

Phosphorylated tau (p-tau) tangles are a neuropathological features of AD, and higher levels of p-tau181 predict faster cognitive decline, even among individuals with normal cognition or mild cognitive impairment (6). Blood-based biomarkers of AD, including p-tau181, have gained significant interest as they are less invasive and more accessible than biomarkers such as cerebrospinal fluid (CSF) and positron emission tomography (PET). (7). Several studies have demonstrated a correlation between blood p-tau levels and tau PET scans and CSF levels of p-tau. Previous studies examining the correlation between AD neuropathological changes and cognitive impairment have shown that the severity of cognitive impairment correlates best with the burden of abnormal tau. Consequently, several clinical trials targeting tau have been conducted and tau-targeted drugs are likely to emerge as future disease-modifying therapies (8,9).

Having an accurate digital biomarker of cognition, such as the ICA, that is correlated with a target neuropathological biomarker can aid in large-scale population screening and regular monitoring during clinical trials. In light of disease-modifying therapies becoming available for AD, the provision of a scalable digital tool to streamline the process of identifying candidates with specific neurodegenerative disorders is promising and beneficial.

While previous studies have shown correlations between the ICA and blood-based biomarkers in MCI and Multiple Sclerosis, its association with p-tau181 remains unknown. This study aims to investigate the relationship between the ICA and blood-based p-tau181 in Alzheimer’s Disease.

## Methods

### Participants

Fifteen mild-AD, twenty three MCI, and twenty two HC participants, including Males and females aged between 50–85 years were recruited to this study. The diagnosis of probable AD was made according to Diagnostic and Statistical Manual Disorders, Fifth Edition (DSM-V), National Institute of Neurological, Communicative Disorders and Stroke-Alzheimer’s Disease and Related Disorder Association (NINCDS-ADRDA) criteria and under the supervision of a geriatric neurologist. All participants provided informed consent in accordance with the Declaration of Helsinki. Tehran University of Medical Sciences ethics committees approved the study (IR.ACECR.ROYAN.REC.1399.059).

Individuals with significant cerebrovascular disease, major psychiatric disorders, repeated head trauma, severe visual impairment, or major medical comorbidities were excluded from the study. Those using cognitive-enhancing drugs or with a history of alcohol or drug abuse were also excluded.

### Study design and data collection

Data collection involved obtaining blood samples and administering two cognitive assessments: the Montreal Cognitive Assessment (MoCA) and the Integrated Cognitive Assessment (ICA).

#### MoCA

MoCA is a ten-minute pen and paper test with a maximum score of 30 to assess visuospatial, memory, attention, and language abilities to detect cognitive impairment in older adults. It is administered by a trained examiner and the results assist in the diagnosis of participants (10). The MoCA has revealed a high sensitivity in the detection of MCI and AD patients (11).

#### Integrated Cognitive Assessment (ICA)

The ICA is a 5-minute, AI-enabled computerized cognitive assessment tool that measures both accuracy and reaction times through a rapid visual categorization task. This tool offers several advantages, including efficient administration, brief duration, automatic scoring, and independence from language and education levels. Its AI capabilities also enhance generalization across diverse populations, making it a robust tool for integration into medical records or research databases.

#### Biomarker Measurement

Participants were asked to have their normal morning routine before the venipuncture. Blood samples were collected into two tubes.

The tube containing silica particles, which activate clotting, were centrifuged at 2000 g for 10 min in 4 °C. Serums were then divided in 4 different microtubes and frozen in -70 °C until further use. Serum samples were analyzed for levels of p-tau181 using 96-well Zellbio ELISA kits based on kits’ protocols (12).

The other tubes contained EDTA as an anticoagulant were stored in 4 °C fried until further use for gene sequencing to determine the specific apolipoprotein E (APOE) genotypes in patients. APOE genotyping was also performed through polymerase chain reaction (13).

### Statistical Analyses

Linear regression models were used to predict biomarker levels in the serum based on ICA index, demographic data, and APOE ε4 status. Cross-validation techniques were employed to assess model performance and significance was set at p < 0.05. All statistical analyses were conducted using Python3.

## Results

### Demographics

Our cross sectional study comprised of participants from three categories. A detailed breakdown of demographic and clinical characteristics is provided in Table 1.

**Table 1.**
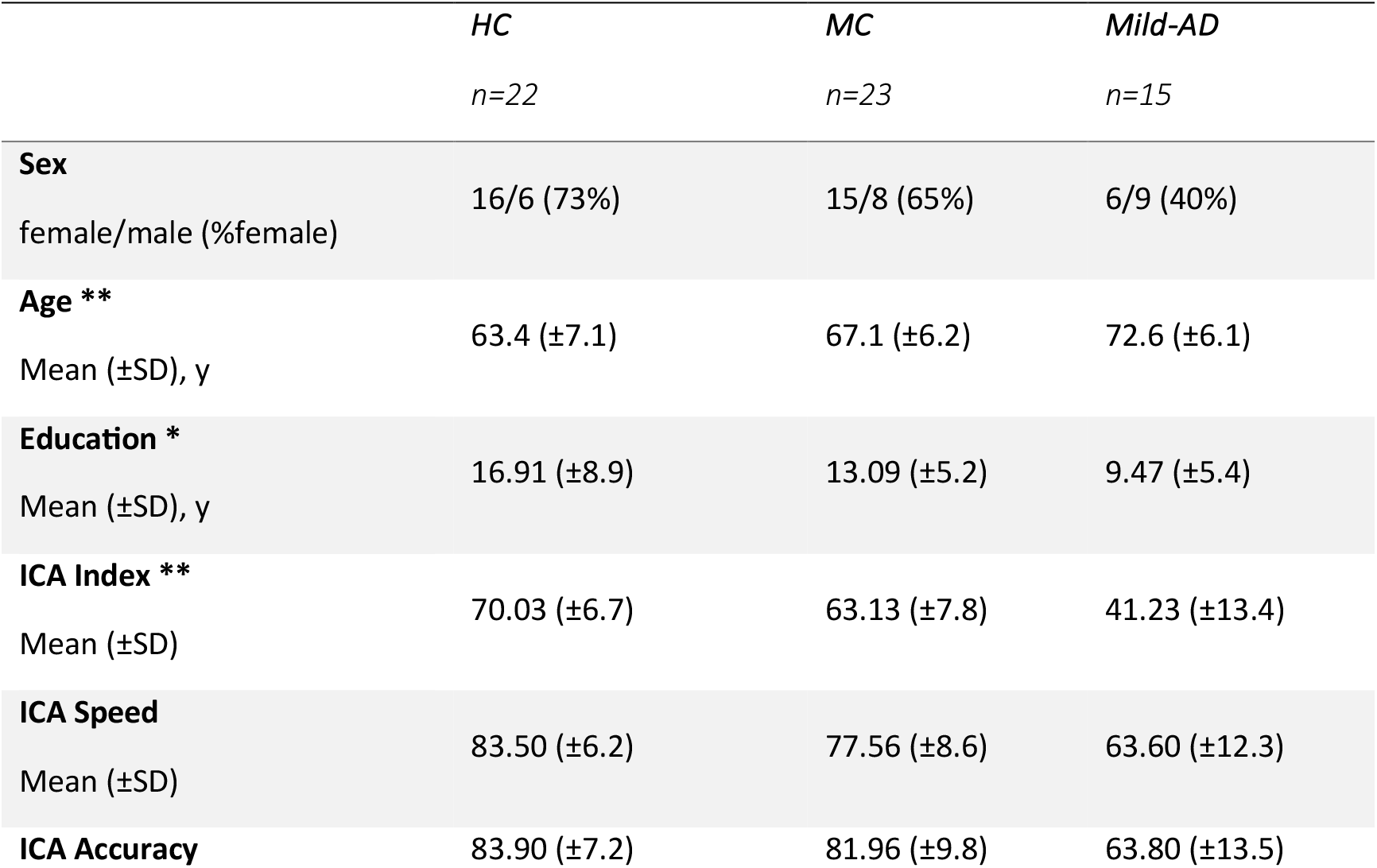
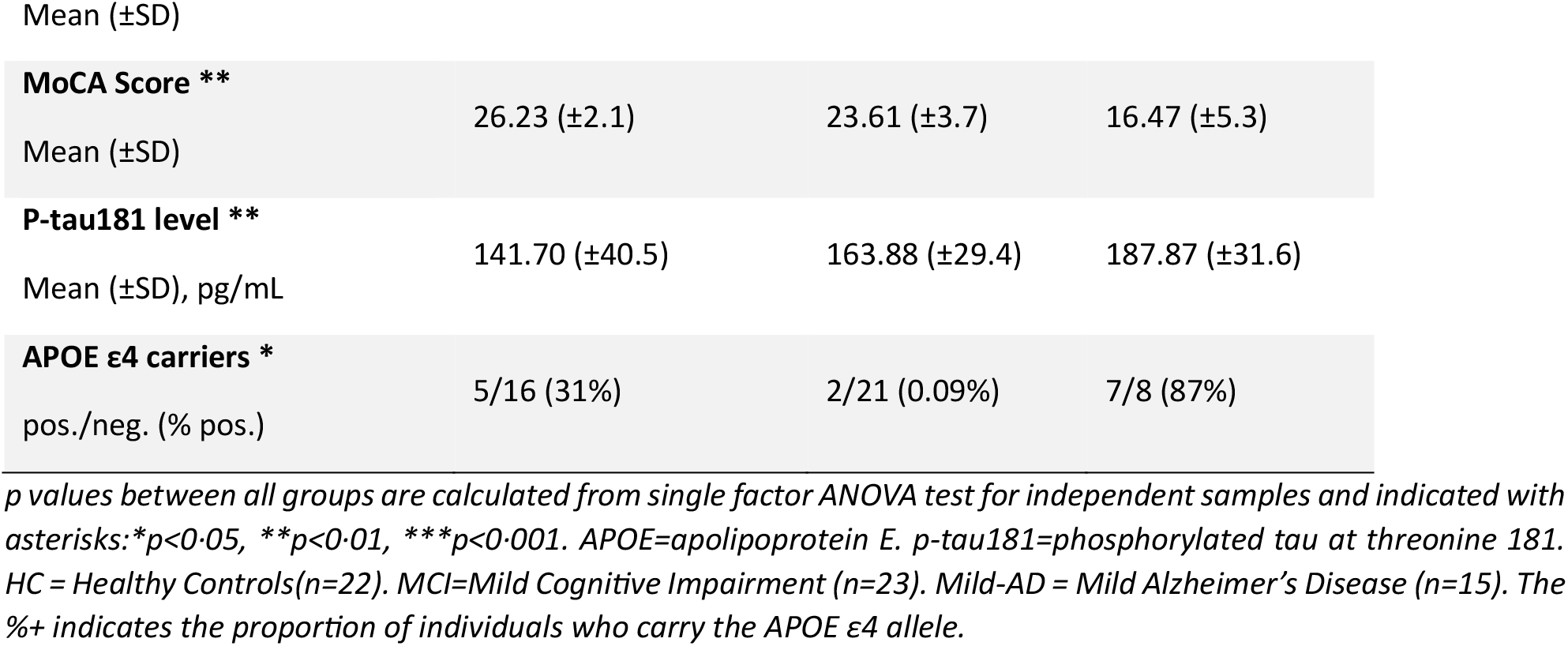
Demographics.

**Table 2.** performance of APOE ε4 carriers and non-carriers in cognitive assessments. A maximum ICA index is 100 and a maximum MoCA score is 30. APOE ε4 -/- indicates the individuals with no allele of ε4. APOE ε4 +/- are the individuals with one allele of ε4.

| Cognitive Assessment | APOE $\epsilon 4$ -/-<br>n = 45 | APOE $\epsilon 4$ +/-<br>n = 14 |
| --- | --- | --- |
| ICA index *<br>mean (±SD) | 62.64 (±11.69) | 51.73 (±20.44) |
| MoCA score **<br>mean (±SD) | 23.80 (±4.68) | 19.57 (±6.28) |

At baseline and irrespective of diagnostic group, p-tau181 level was positively associated with age (r, 0.36; P < .001) but not with educational level or sex.

#### Distribution of ICA index across diagnostic groups

The distribution of the ICA index across diagnostic groups (Figure 1) showed a significant difference between healthy controls, mild cognitive impairment, and mild Alzheimer’s disease. The ICA was found to be a sensitive tool in discriminating between these groups, with a p-value of less than 0.001.

**Figure 1.**
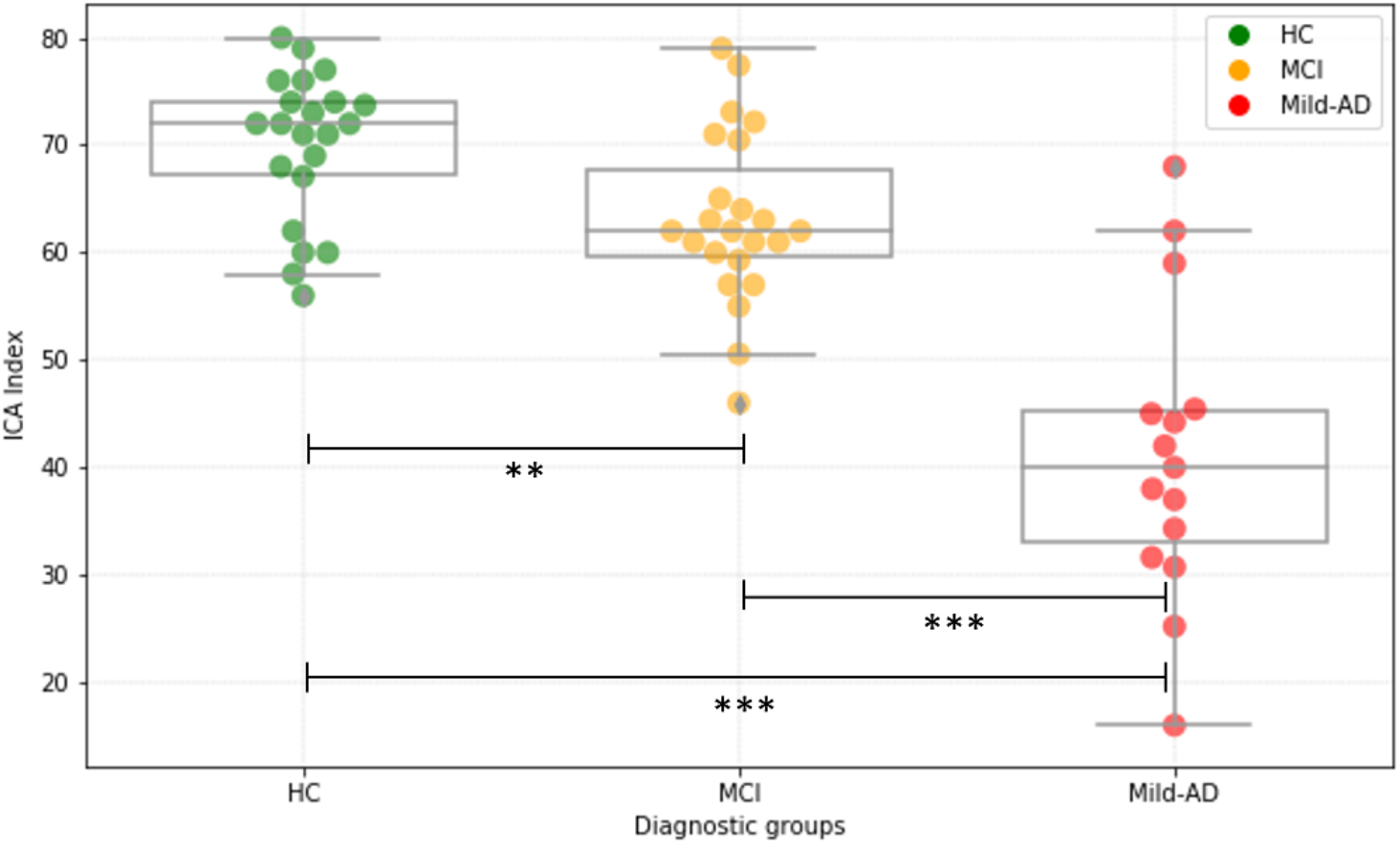
Distribution of ICA index across diagnostic groups. Each dot indicates an individual (n= 60). Boxes show interquartile range and the horizontal lines are medians. p values are indicated with asterisks:^*^p<0·05, ^**^p<0·01, ^***^p<0·001. HC = Healthy Controls (n=22). MCI=Mild Cognitive Impairment (n=23). Mild-AD = Mild Alzheimer’s Disease (n=15).

#### Serum p-tau181 concentration across diagnostic groups

The serum p-tau181 concentration also showed a significant difference between the three diagnostic groups, with the highest levels seen in the mild Alzheimer’s group. The p-value for the difference between healthy controls and mild Alzheimer’s disease was less than 0.001, and between mild cognitive impairment and mild Alzheimer’s disease was less than 0.05 (Figure 2).

**Figure 2.**
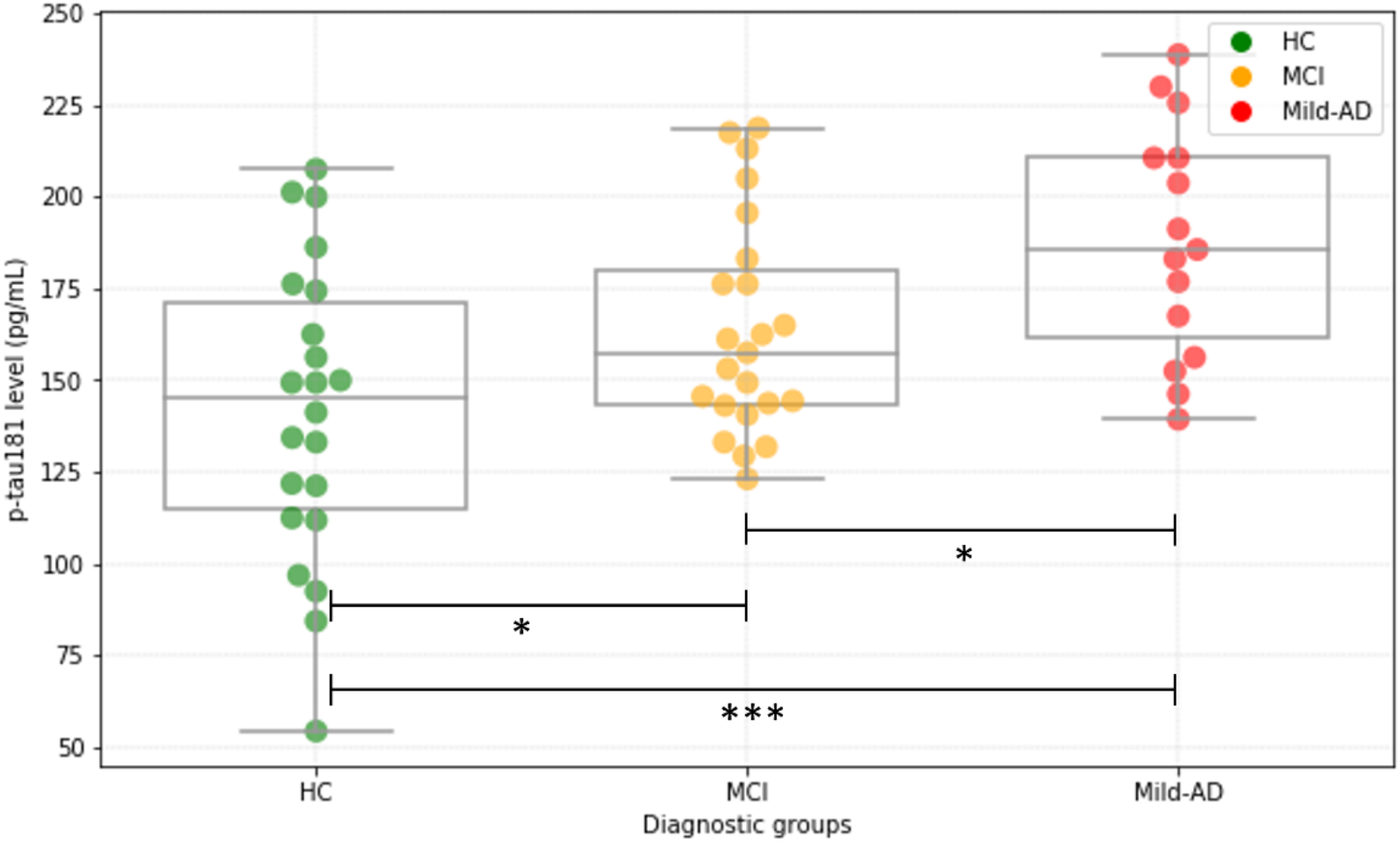
Serum p-tau181 concentration across diagnostic groups. Each dot indicates an individual (n= 60). Boxes show interquartile range and the horizontal lines are medians. p values are indicated with asterisks:^*^p<0·05, ^**^p<0·01, ^***^p<0·001. HC = Healthy Controls(n=22). MCI=Mild Cognitive Impairment (n=23). Mild-AD = Mild Alzheimer’s Disease (n=15). p-tau181=tau phosphorylated at threonine 181.

### Relationship between ICA Index and serum p-tau181

A scatter plot of p-tau181 level against ICA Index (Figure 3) reveals a negative correlation, indicating that as the ICA Index increases, p-tau181 level tends to decrease. The Spearman correlation coefficient is −0.35 (p=0.006).

**Figure 3.**
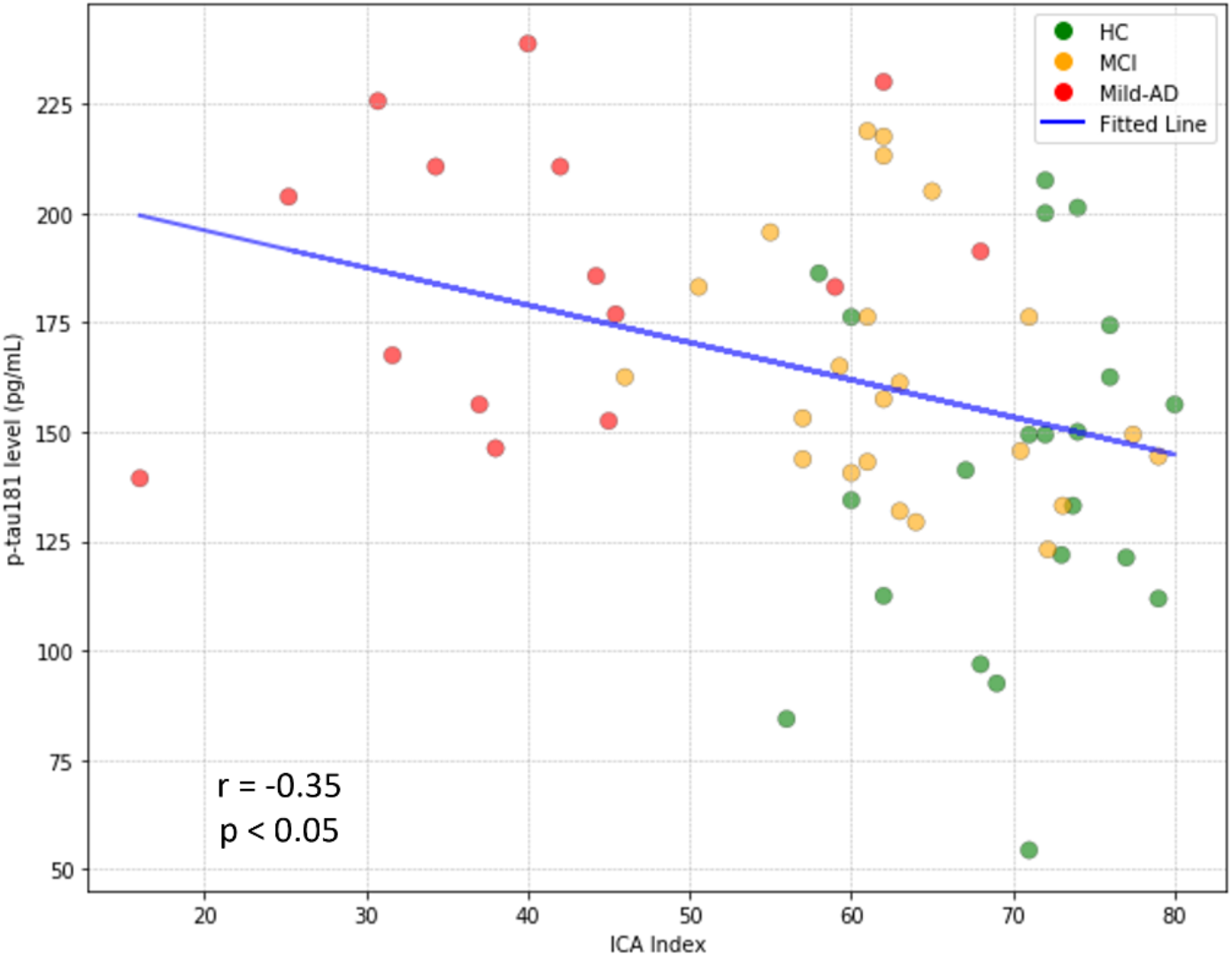
Scatter plot of p-tau181 level against ICA index. Individuals are color-coded by their categories. The blue line represents the linear regression fit. HC = Healthy Controls (n=22). MCI=Mild Cognitive Impairment (n=23). Mild-AD = Mild Alzheimer’s Disease (n=15). p-tau181=tau phosphorylated at threonine 181.

### Relationship between MoCA score and serum p-tau181

Similarly, a scatter plot of p-tau181 level against MoCA score (Figure 4) showed a negative correlation, with a Spearman correlation coefficient of -0.28 (p=0.032). This suggests that as cognitive function declines, p-tau181 levels tend to increase.

**Figure 4.**
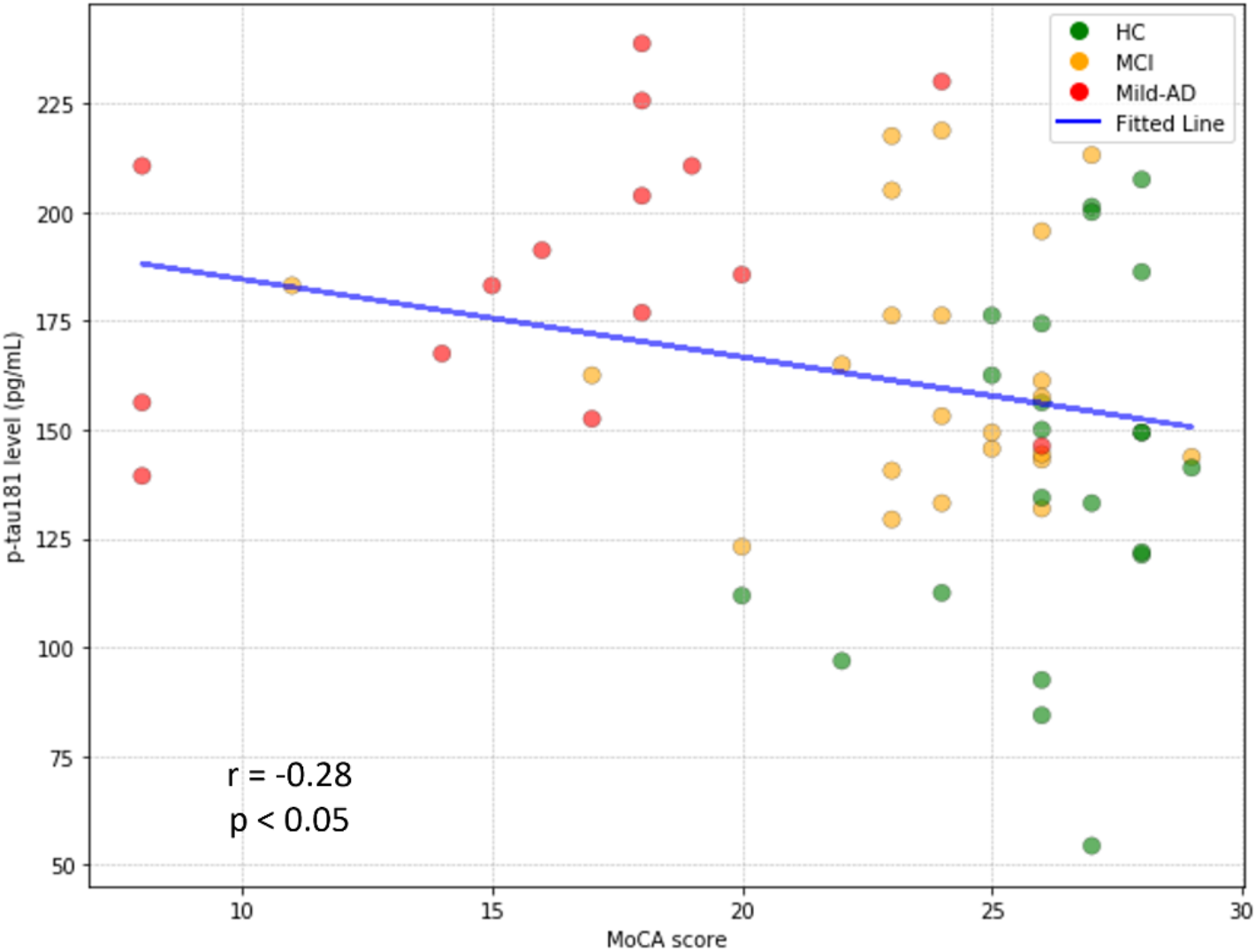
Scatter plot of p-tau181 level against MoCA score. Individuals are color-coded by their categories. The blue line represents the linear regression fit. HC = Healthy Controls(n=22). MCI=mild cognitive impairment (n=23). Mild-AD = Mild Alzheimer’s Disease (n=15). p-tau181=tau phosphorylated at threonine 181.

### Comparison of explanatory power: ICA Index vs. MoCA score

Both ICA index and MoCA score show a negative correlation with p-tau181 level, meaning that as the variable increases, p-tau181 level tends to decrease. However, ICA index has a slightly stronger negative correlation with p-tau181 level compared to MoCA score, as evidenced by the larger magnitude of the correlation coefficients (Figure3 and Figure4).

In a nested model comparison, the ICA Index (Spearman’s R = -0.35) did not significantly better explain the variance in p-tau181 level compared to MoCA score (Spearman’s R = -0.28). The ICA Index accounted for approximately 10.77% of the variance in p-tau181 level, while MoCA score explained about 6.15%.

### Relationship between cognitive assessments and APOE status

The analysis of the relationship between cognitive assessments (ICA index and MoCA score) and APOE ε4 status (Figure 5) showed that APOE ε4 carriers had lower scores on both the ICA index and MoCA score compared to non-carriers, with p-values of less than 0.05. However, there was no significant difference in serum p-tau181 levels between APOE ε4 carriers and non-carriers.

**Figure 5.**
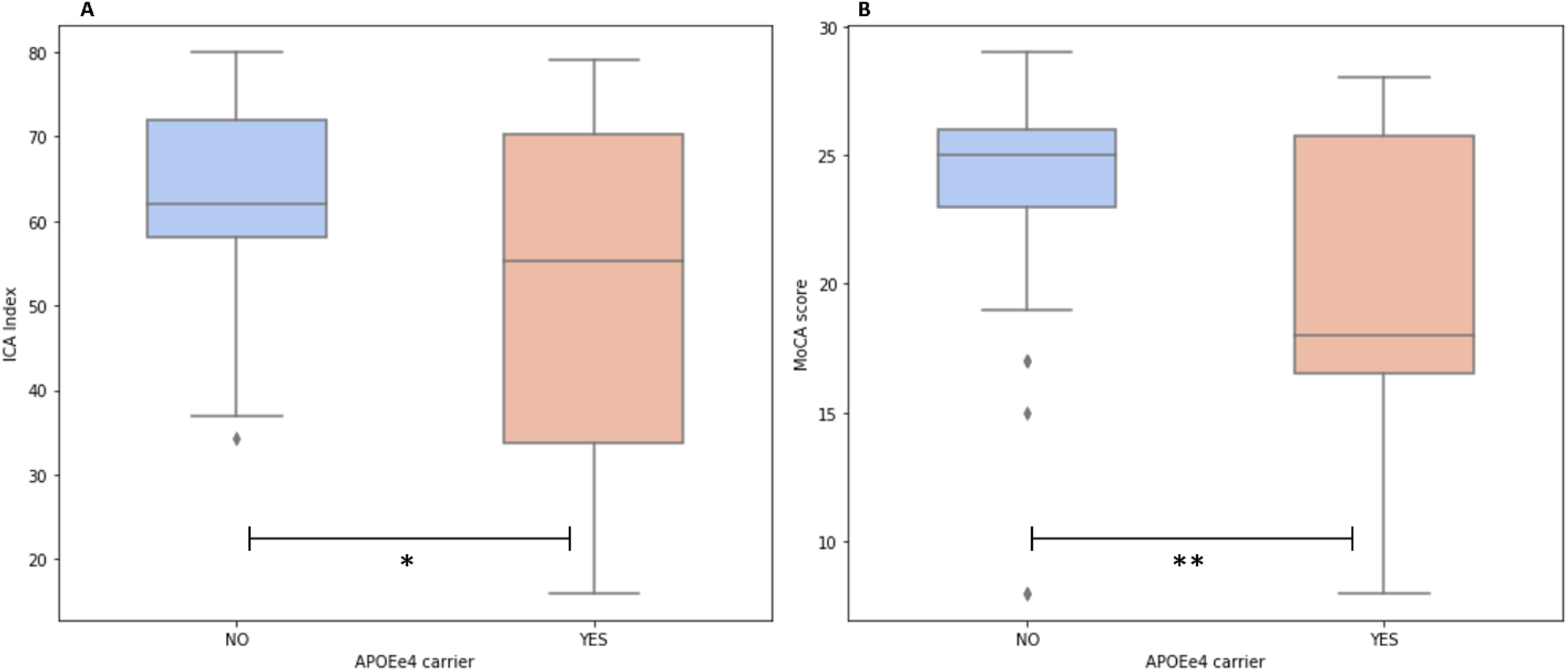
Cognitive performance in APOE ε4 carriers and non-carriers. **A**, ICA index in APOE ε4 carriers and non-carriers. **B**, MoCA score in APOE ε4 carriers and non-carriers. Boxes show interquartile range and the horizontal lines are medians. Significant p values are indicated with asterisks:^*^p<0·05, ^**^p<0·01.

### An ICA model to predict biomarkers level in the serum

A combination of the ICA, demographic data, and APOE ε4 status was used to predict the level of biomarkers in the serum (Figure 6). An example model achieved a coefficient of determination of 0.06 in predicting p-tau181 levels.

**Figure 6.**
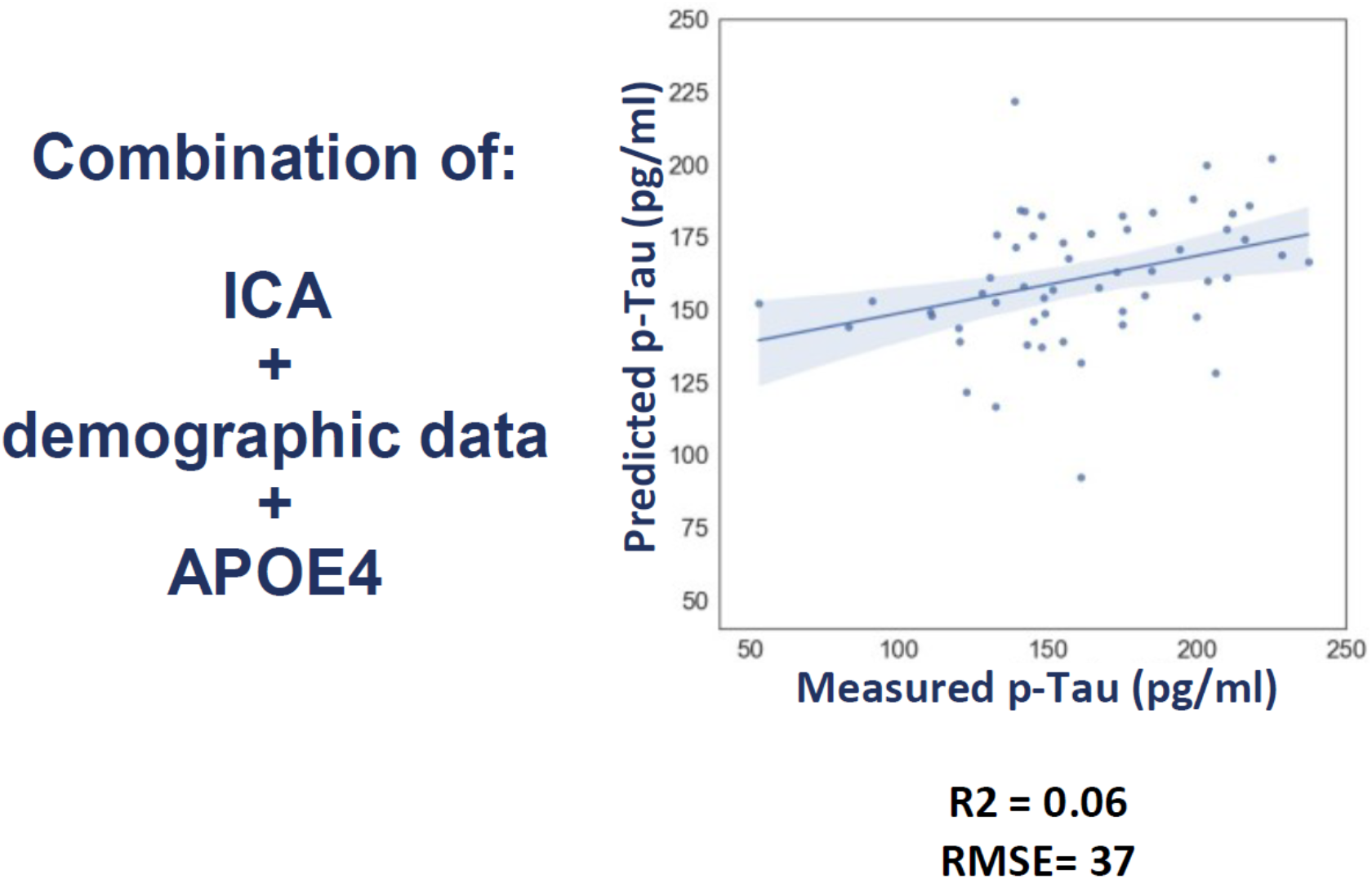
Linear regression model predicting serum p-tau181 levels using a combination of the Integrated Cognitive Assessment (ICA), demographic data, and APOE ε4 status. The model achieved a coefficient of determination (R^2^) of 0.06, indicating a modest level of explained variance in p-tau181 levels. The model’s prediction performance is further indicated by a Root Mean Square Error (RMSE) of 37 and a Pearson correlation coefficient (r) of 0.34 (p=0.01), suggesting a statistically significant but moderate correlation between the predicted and measured p-tau181 values.

## Discussion

This study explored the association between the Integrated Cognitive Assessment (ICA), a computerized self-administered cognitive tool, and serum p-tau181 levels across healthy controls, mild cognitive impairment (MCI), and mild Alzheimer’s Disease (AD) groups. The ICA was found to significantly differentiate between these diagnostic groups and exhibited a significant association with elevated levels of p-tau181. These findings suggest the potential utility of ICA as a scalable and cost-effective tool for AD screening and monitoring.

Our study aligns with previous research that has investigated the correlation of ICA with serum biomarkers. In previous studies, we observed a significant correlation between ICA and serum neurofilament light (NfL) levels in multiple sclerosis (14). Another study comparing healthy controls with patients with MCI showed that the neural speed of animacy information processing is decreased in MCI patients. They suggested that the speed and pattern of animacy information processing provide clinically useful information as a potential biomarker for detecting early changes in MCI and AD patients (15).

Both the ICA and MoCA showed associations with serum p-tau181 levels, a biomarker linked to cognitive decline in AD. The ICA’s rapid visual categorization task may enhance its sensitivity to early cognitive changes, particularly in less severe stages of brain deterioration. This sensitivity could make ICA a valuable tool for large-scale screening to identify individuals at risk for MCI and early-stage AD. Our study supports the notion that p-tau181 is a specific biomarker for AD rather than a general dementia biomarker, with elevated levels observed in participants with mild AD compared to those with MCI and healthy controls (2,16).

The presence of the APOE ε4 allele was significantly associated with cognitive decline as measured by both ICA and MoCA, reinforcing its role as a genetic risk factor for late-onset AD (17). However, no significant association was found between APOE ε4 status and serum p-tau181 levels. This suggests that while APOE ε4 may play a significant role in cognitive decline, its influence on tau pathology might be indirect or mediated by other factors.

In addition to p-tau181, the study also measured the serum Aβ42/40 ratio but found no significant correlation with either the diagnostic groups or the ICA index(18). This inconclusive correlation could be due to the use of the less sensitive ELISA method compared to more advanced assays like Quantrix (19). Nevertheless, the lack of a strong association underscores the complexity of AD pathology and the need for multiple biomarkers in the diagnostic process.

The ICA shows promise as a tool for large-scale cognitive screening, particularly in the context of identifying candidates for p-tau181-targeted drug trials and disease-modifying therapies. Its scalability, ease of administration, and potential to detect early cognitive changes make it a suitable option for population-wide screening initiatives. Moreover, by integrating cognitive assessment with blood-based biomarkers, ICA could aid in the early detection and monitoring of AD, contributing to improved management strategies.

### Strengths and Limitations

This study’s strengths include the implementation of the ICA as a scalable and accessible cognitive assessment tool and the use of a relatively affordable method for measuring serum p-tau181 levels. However, the study’s limitations include a relatively small sample size and a cross-sectional design, which restricts the ability to establish causality or track longitudinal changes. The absence of tau PET scans at the time of plasma p-tau181 measurement is another limitation, as such imaging could have provided complementary information about tau pathology.

### Future work

Future studies should focus on larger, more diverse populations and adopt longitudinal designs to validate the findings and explore the progression of cognitive decline in relation to serum biomarkers. Expanding biomarker assessments to include amyloid beta, glial fibrillary acidic protein (GFAP), and neurofilament light (NfL) could provide a more comprehensive understanding of AD pathology. Integrating advanced imaging techniques like tau PET scans alongside plasma biomarker measurements could offer deeper insights into the disease process.

## Data Availability

All data produced in the present study are available upon reasonable request to the authors

## Supplementary information

## Acknowledgements

I would like to express my heartfelt gratitude to all those who supported me throughout my research journey. I am especially grateful to Mahdieh Khanbagi and Hamed Karimi for their support and great help in patients recruitment and data management.

## Ethics approval and consent to participate

The study was conducted according to the Declaration of Helsinki and approved by the Royan Institute’s ethics committee (IR.ACECR.ROYAN.REC.1399.059). Informed written consent was obtained.

## Supplement

Clinical trial registration available on https://www.isrctn.com/ISRCTN18112405.

### Pairwise Relationships between ICA Metrics

The analysis of the pairwise relationships between ICA metrics shows a strong positive correlation between the ICA Index and both ICA Accuracy and ICA Speed. Given that the ICA Index is composed of these two components, this result is expected. However, what is particularly noteworthy in this population is the balance between speed and accuracy, as indicated by the moderate correlation between these two metrics (Spearman’s R = 0.27). This suggests that individuals in this cohort have not sacrificed one aspect— speed or accuracy—in favor of the other. Instead, there appears to be a harmonious relationship where improvements in one dimension are not achieved at the expense of the other. This balance may reflect a well-rounded cognitive performance within this specific population.

**Supplementary Figure 2.**
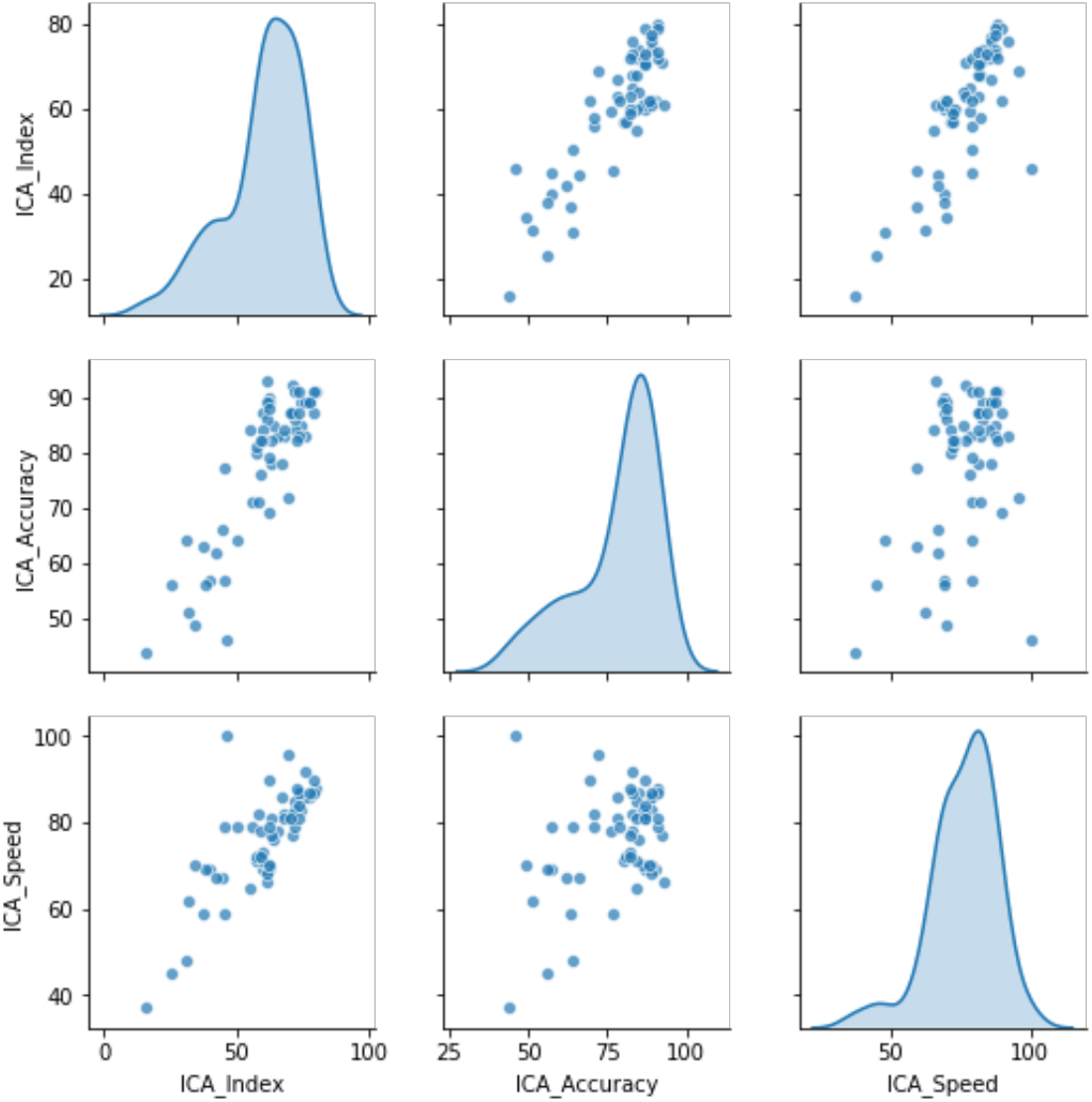
The scatter plots visualize the relationships between the ICA index, accuracy, and Speed. The scatter plots illustrate the relationships among the ICA Index, ICA Accuracy, and ICA Speed. The plots demonstrate a strong positive correlation between the ICA Index and both ICA Accuracy and ICA Speed, with Spearman’s R values of 0.76 and 0.78, respectively. A moderate positive correlation is also observed between ICA Speed and ICA Accuracy (Spearman’s R = 0.27).

### APOE status and its relationship with serum p-tau181

The distribution of p-tau181 levels across APOE ε4 carriers and non-carriers showed no significant difference between the two groups.

**Supplementary Figure 2.**
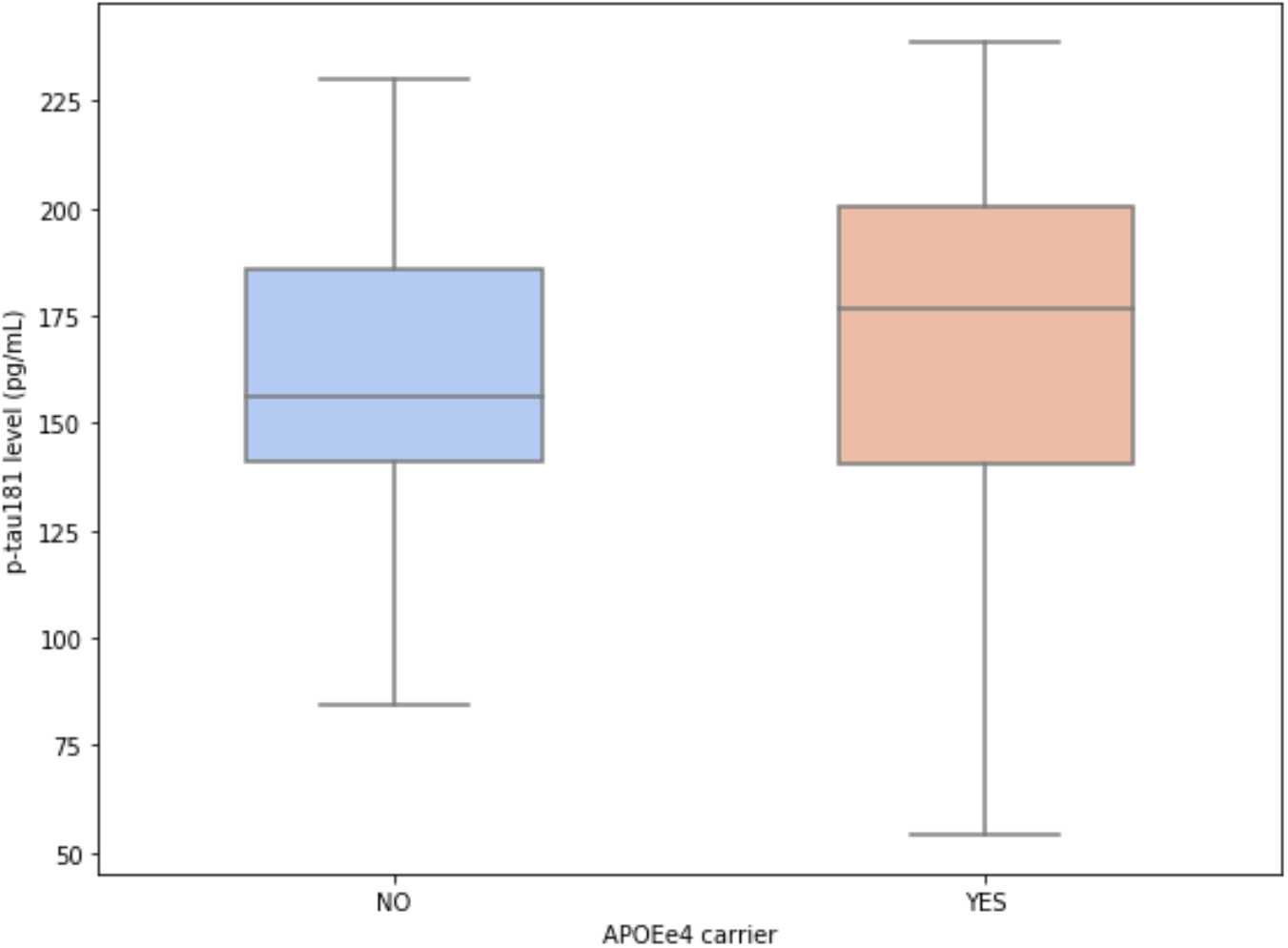
Box plots representing the distribution of p-tau181 level across APOE ε4 carriers and non-carriers categories. The p value of the difference between groups is not significant.

Mean(±SD) for carriers: 168.03 (±48.67), Mean(±SD) for non-carriers : 161.24 (±34.03)

